# A RCT of a third dose CoronaVac or BNT162b2 vaccine in adults with two doses of CoronaVac

**DOI:** 10.1101/2021.11.02.21265843

**Authors:** Chris Ka Pun Mok, Samuel M.S. Cheng, Chunke Chen, Karen Yiu, Tat-On Chan, Kiu Cheung Lai, Kwun Cheung Ling, Yuanxin Sun, Lun Lai Ho, Malik Peiris, David S Hui

## Abstract

**Background:** Poor immunogenicity and antibody waning were found in vaccinees of CoronaVac. There is lack of randomized controlled trial (RCT) data to compare the immunogenicity and safety of schedules using homologous and heterologous vaccine as a booster dose.

**Methods:** We randomly assigned adults who had received 2 doses of CoronaVac with low antibody response to receive an additional booster dose of either BNT162b2 or CoronaVac. The local and systemic adverse reactions were recorded. Levels of SARS-CoV-2 neutralizing and spike binding antibody in plasma were measured.

**Findings:** At one month after the third dose of vaccine, BNT162b2 vaccines elicited significantly higher surrogate virus neutralizing test (sVNT), spike receptor binding, spike N terminal domain binding, spike S2 domain binding levels than CoronaVac. More participants from the BNT162b2 group reported injection site pain and swelling as well as fatigue and muscle pain than those who received CoronaVac as the third dose. The mean results of the sVNT against the wild type, beta, gamma and delta variants in the BNT162b2 boosted group was 96.83%, 92.29%, 92.51% and 95.33% respectively which were significantly higher than the CoronaVac boosted group (Wild type: 57.75%; Beta: 38.79 %; Gamma: 32.22%; Delta: 48.87%)

**Conclusion:** Our RCT study shows that BNT162b2 booster dose for those people who poorly responded to the previous vaccination of CoronaVac is significantly more immunogenic than a CoronaVac booster. BNT162b2 also elicits higher levels of SARS-CoV-2 specific neutralizing antibodies to different variants of concern. The adverse reactions were only mild and short-lived.

**At a Glance Commentary:** *Scientific Knowledge on the Subject:* Poor immunogenicity and antibody waning were found in vaccinees of CoronaVac. There is lack of randomized controlled trial (RCT) data to compare the immunogenicity and safety of schedules using homologous and heterologous vaccine as a booster dose.

*What This Study Adds to the Field:* Our RCT study shows that BNT162b2 booster dose for those people who poorly responded to the previous vaccination of CoronaVac is significantly more immunogenic than a CoronaVac booster. The adverse reactions were only mild and short-lived.

## Introduction

Over two hundred million cases and over 50 million deaths have been reported since the emergence of the COVID-19 pandemic in late 2019 (1). So far, seven Covid-19 vaccines have received Emergency Use Listing by the World Health Organization (WHO) and two of them are adjuvanted inactivated virus vaccines (2). CoronaVac is one of the WHO approved inactivated virus vaccines and over 750 million doses have been administered in more than 40 countries. The safety and performance of this vaccine have been evaluated in different age groups (3-5). The phase three randomized clinical trials (RCT) of CoronaVac showed efficacies against symptomatic illness of 50.65%, 65.30%, 83.50% in Brazil, Indonesia and Turkey respectively (6). Importantly, the protection for hospitalization and death appeared even higher in these studies (7,8). However, the recent breakthrough infections including severe disease and death in CoronaVac vaccinated adults in Indonesia has raised concern on the effectiveness of CoronaVac (9). Our recent observational study has shown that the immunogenicity of CoronaVac is much lower compared to the BNT162b2 mRNA vaccine (10). From plaque reduction neutralization test titres, we assessed that most vaccine recipients reached protective thresholds at one month after the second dose of vaccine but many would fall below protective levels within a few months allowing for an inevitable two-fold waning of antibody titers (10). This protection may be further compromised by virus variants that are known to reduce vaccine protection (11). Thus, a booster dose of vaccine would be desirable. However, the question of whether a homologous or heterologous vaccine should be used as the booster dose is not yet clear. A head-to-head comparison between the two approaches in an unbiased experimental setting is thus urgently needed. Here, we report the results of a RCT to compare the immunogenicity and safety of using BNT162b2 and CoronaVac as a booster dose for adults with low antibody response to two doses of CoronaVac.

## Materials and Methods

### Cohort study design and participants

376 healthy adults aged between 19-77 years old who had received two doses of CoronaVac were recruited for our previous study in in Hong Kong SAR, China at the community vaccination centers at the Chinese University of Hong Kong Medical Centre, Prince of Wales Hospital and Kowloon Bay Vaccination Station between March 10 and August 31, 2021 (10). The percentage (%) inhibition of binding of SARS-CoV-2 spike receptor binding domain (RBD) to the ACE-2 receptor was tested by surrogate virus neutralization test (sVNT) in the plasma of 360 participants (10). Based on the previous study of convalescent antibody titers of RT-PCR confirmed convalescent symptomatic patients, we have estimated that the 20% convalescent antibody titer threshold for 50% protection from re-infection for PRNT_50_ was 1:25.9 (95% CI 1:24.7-1:27.6) (12). Since there will be waning of antibody from the peak titres observed at 1-month post second dose of vaccine, we set the target titre to be achieved at 1 month post-second dose of vaccine to be twice the 50% protection titre, i.e 1:52. This corresponds to a sVNT inhibition of 60% (Supplementary Figure 1) (10, 12). 230 out of 360 in our cohort showed the sVNT results below 60% in their plasma specimens which were collected at one month after the second dose. Eighty participants of age between 34-73 years old were invited to join our third dose RCT between August and October, 2021. The participants were randomized to receive either BNT162b2 (n=40) or CoronaVac (n=40) as the third dose. Safety assessment objectives included recording of local or systemic reactogenicity events from each participant at 1 week of vaccination. Ten ml of heparinized blood were collected from each donor before vaccination and at one month after receiving the third dose of either BNT162b2 or CoronaVac.

### Study approval

The study was approved by the Joint Chinese University of Hong Kong-New Territories East Cluster Clinical Research Ethics Committee (Ref no: 2020.229) and all participants provided written consent. This clinical trial has been registered at ClinicalTrials.gov with identifier NCT04611243

### Processing and storage of specimens

Heparinised blood samples were collected and centrifuged at 3000g for 10 minutes at room temperature. The plasma was collected and stored at -80C.

### Surrogate virus neutralization test (sVNT)

SARS-CoV-2 surrogate virus neutralization test kits and the RBD protein of beta, gamma and delta varients tagged with HRP were obtained from GenScript, Inc., NJ, USA, and the tests were carried out according to the manufacturer’s instructions and our previous study (10). The assay validity was based on values representing optical density at 450 nm (OD450) for positive and negative results falling within the range of recommended values. On the basis of the assumption that the positive and negative controls gave the recommended OD_450_ values, percent inhibition of each serum was calculated as follows: percent inhibition (1 - sample OD value/negative-control OD value) x 100. Percent inhibition values of 30% or more are regarded as positive results according to the manufacturer’s recommendation.

### ELISA for spike RBD, NTD, and S2 antibodies

Each human plasma sample was diluted to 1:100 in Chonblock blocking/sample dilution buffer and the ELISA was performed according to our previous protocol (13). The RBD and NTD protein were produced according to the previous study (14) and S2 protein were purchased from Sino Biological (China).

### Outcomes

The primary outcomes were humoral immunogenicity were measured by sVNT, PRNT and ELISA in the plasma samples collected at one month after the third dose of vaccination. The secondary outcome was the occurrence of adverse reactions within 7 days and 1 month after the third dose of vaccination.

### Sample size calculation

The sample size was calculated based on the results of our previous study (10). The standard deviation (SD) of % inhibition in the sVNT test in the post-vaccine plasma from our age-matched cohort for BNT1626 and CoronaVac were 3.45 and 16.72 respectively. A sample size of 32 patients in each group was estimated to have 90% power to detect a difference of 10% in sVNT by using a two-sided, unpaired t-test. As the antibody response after the 3rd dose is unclear, we have chosen a conservative sample size of 40 in each group.

### Statistical analysis

Continuous variables were summarized as mean with SD while categorical variables were summarized as frequency. Geometric means were used for comparison of neutralization titers. Comparison between groups was conducted with two-sided, unpaired t-test for continuous variables and the Chi-square test or Fisher’s exact test for categorical variables as appropriate. All analyses were performed in R (version 4.1.0; R Foundation for Statistical Computing) or Prism 9 (Graphpad). P values < 0.05 were considered statistically significant.

## Results

We carried out a RCT of the third dose vaccination between August 18 and October 26, 2021 on the community subjects who had received two doses of CoronaVac but shown low immune response against SARS-CoV-2. The subjects were recruited from the cohort of our previous observational study who had received two doses of CoronaVac (10). We observed that 230 out of 360 (63.89%) individuals who had received two doses of CoronaVac showed sVNT results lower than 60% of inhibition. 80 out of the 230 “low responders” were enrolled for this RCT study and were randomized to receive an additional dose of either BNT162b2 (n=40) or CoronaVac (n=40). The study design is summarized in Figure 1.

**Figure 1.**
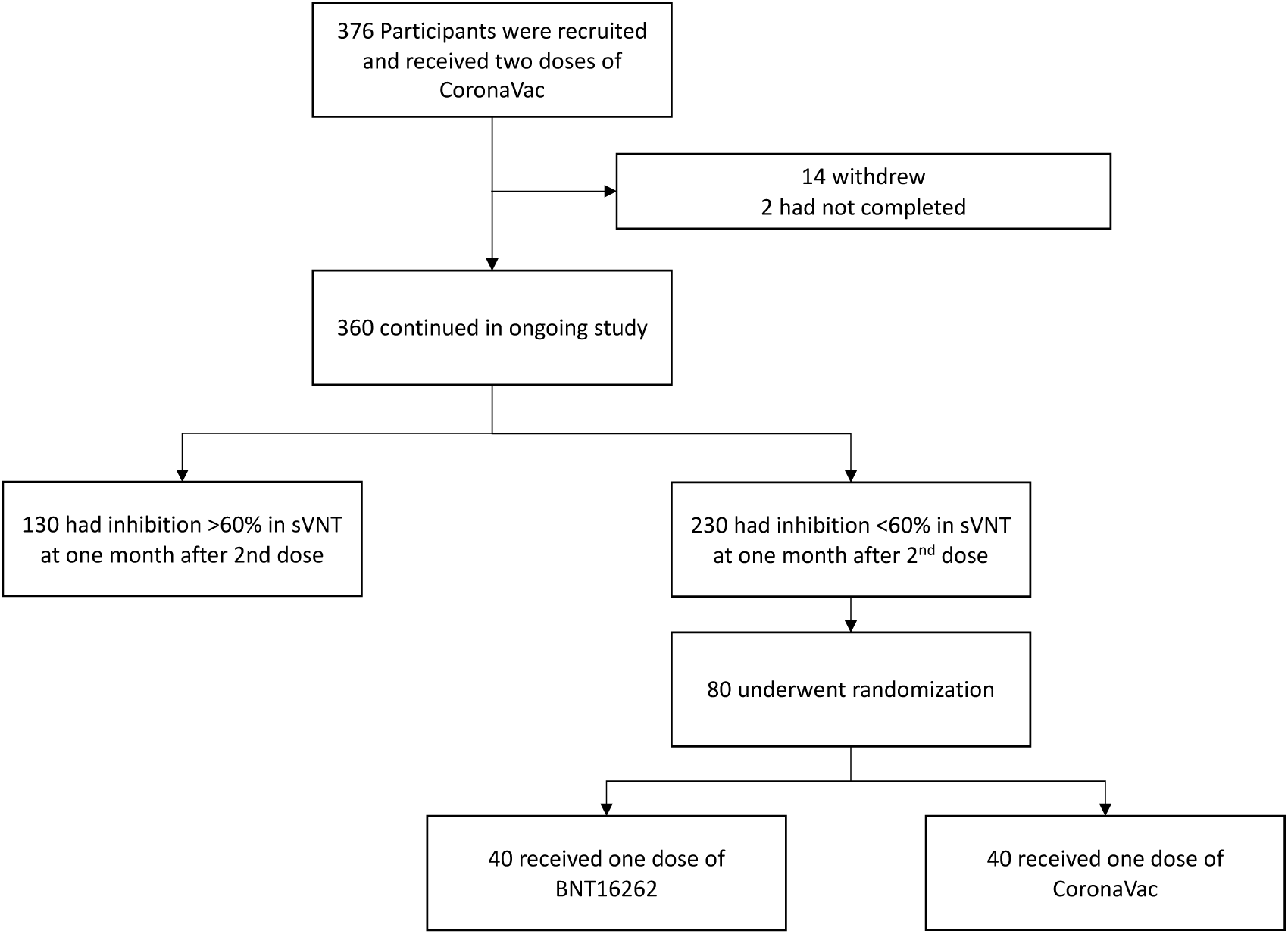
Flow chart of the randomized clinical trial study.

All participants were adults within the age-range 34 to 73 years. The mean (SD) age of the BNT162b2 (3^rd^ dose) and CoronaVac (3^rd^ dose) groups were 51.20 (8.79) and 51.50 (8.83) years respectively. The age (p=0.883) and gender (p=0.482) did not significantly differ between the two groups. Other demographic information, including comorbidities, the habits of smoking, alcohol use, history of other vaccinations and frequency of exercise were also not significantly different. The mean duration between the second and the third dose of the two groups were 112.28 and 115.95 days respectively (p=0.590) (Table 1).

**Table 1.**
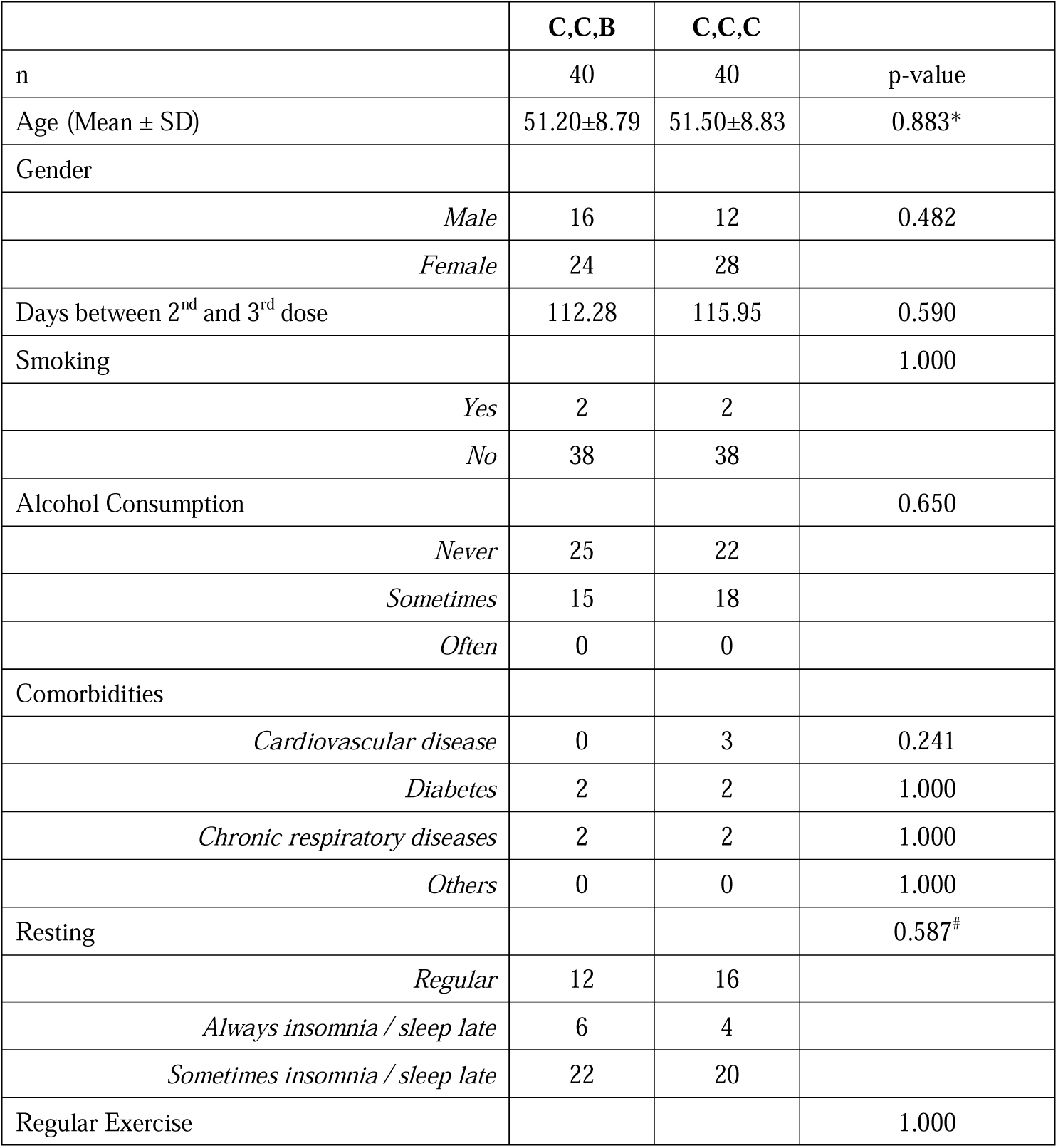
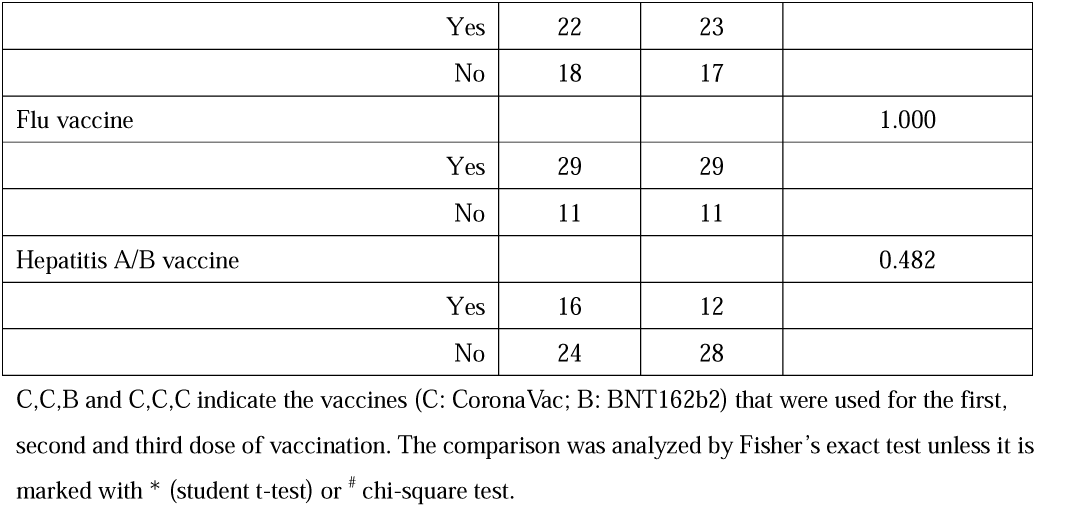
Demographic Characteristics of the Participants.

The local and systemic adverse reactions were assessed and compared between the two groups (Table 2). More participants in the BNT162b2 (3^rd^ dose) group reported pain (p<0.001) and swelling (p<0.05) at the injection site than those receiving CoronaVac as the third dose. There were significantly more participants in the BNT162b2 (3^rd^ dose) group that complained of fatigue (p<0.01) and muscle pain (p<0.05) compared to the CoronaVac (3^rd^ dose) group. However, none of these side effects were considered unacceptable by the participants and none of them actively complained about the local or systemic adverse reaction 1 month after the third dose of vaccination.

**Table 2.** Adverse reactions after receiving the third dose of vaccination.

|  | After 2<sup>nd</sup> dose |  |  | After 3<sup>rd</sup> dose |  |  |
| --- | --- | --- | --- | --- | --- | --- |
|  | C,C,B | C,C,C |  | C,C,B | C,C,C |  |
| n | 40 | 40 | p-value | 40 | 40 | p-value |
| **Local reactions** |  |  |  |  |  |  |
| Pain | 12 | 13 | 1.000 | 34 | 12 | <0.001 |
| Erythema | - | - | N.A. | 2 | 0 | 0.494 |
| Pruritus | - | - | N.A. | 3 | 1 | 0.616 |
| Swelling | - | - | N.A. | 14 | 4 | 0.014 |
| **Systemic reactions** |  |  |  |  |  |  |
| Fever* | 2 | 2 | 1.000 | 7 | 1 | 0.057 |
| Fatigue | 16 | 14 | 0.818 | 24 | 10 | 0.003 |
| Diarrhoea | 2 | 3 | 1.000 | 1 | 0 | 1 |
| Muscle pain | 8 | 4 | 0.348 | 13 | 4 | 0.027 |
| Nausea | - | - | N.A. | 2 | 0 | 0.494 |
| Headache | 5 | 5 | 1.000 | 10 | 3 | 0.067 |
| Cough | 2 | 0 | 0.494 | 2 | 2 | 1 |
| Anorexia | 0 | 1 | 1.000 | 4 | 1 | 0.359 |
| Hypoesthesia | - | - | 0.116 | 4 | 0 | 0.116 |
| Dizziness | - | - | 0.264 | 6 | 2 | 0.264 |
| Abdominal distention | - | - | N.A. | 1 | 0 | 1 |
| Peripheral oedema | - | - | N.A. | 1 | 0 | 1 |
| Abdominal pain | - | - | N.A. | 1 | 0 | 1 |
| Vomiting | 0 | 0 | N.A. | 0 | 0 | N.A. |
| Drowsiness | - | - | N.A. | *11* | *8* | 0.601 |
| Joint pains | - | - | N.A. | *6* | *3* | 0.482 |
| Rash | - | - | N.A. | *2* | *0* | 0.494 |
| Palpitation | - | - | N.A. | *5* | *2* | 0.432 |
| Claimed no adverse effect | *15* | *14* | 1.000 | *8* | *16* | 0.087 |

C,C,B and C,C,C indicate the vaccines (C: CoronaVac; B: BNT162b2) that were used for the first, second and third dose of vaccination. \*Oral temperature above 37.5°C was considered as fever. -: Did not included in the questionnaire. N.A.: Not available. The comparison was analyzed by Fisher's exact test.

We used sVNT, which has high overall correlation with the PRNT_50_ titres (11) to examine the level of SARS-CoV-2 specific neutralizing antibody from the plasma samples collected before and at 1 month after the third dose of vaccination. The plasma samples from all vaccinees in both groups were negative in sVNT before any vaccination and the sVNT showed comparable results 1 month after the second doses as expected (Figure 2A). Only 8 and 9 participants still showed sVNT inhibition higher than 30% before they received the BNT162b2 and CoronaVac as the third dose respectively. One month after the third dose of vaccination, the mean % inhibition in the sVNT test in the plasma for the BNT1626 and CoronaVac groups was 96.83% (SD 2.74) and 57.75% (SD 24.68), respectively (p<0.0001) (Figure 2A). Only five participants boosted with CoronaVac showed % of inhibition higher than 90%.

**Figure 2.**
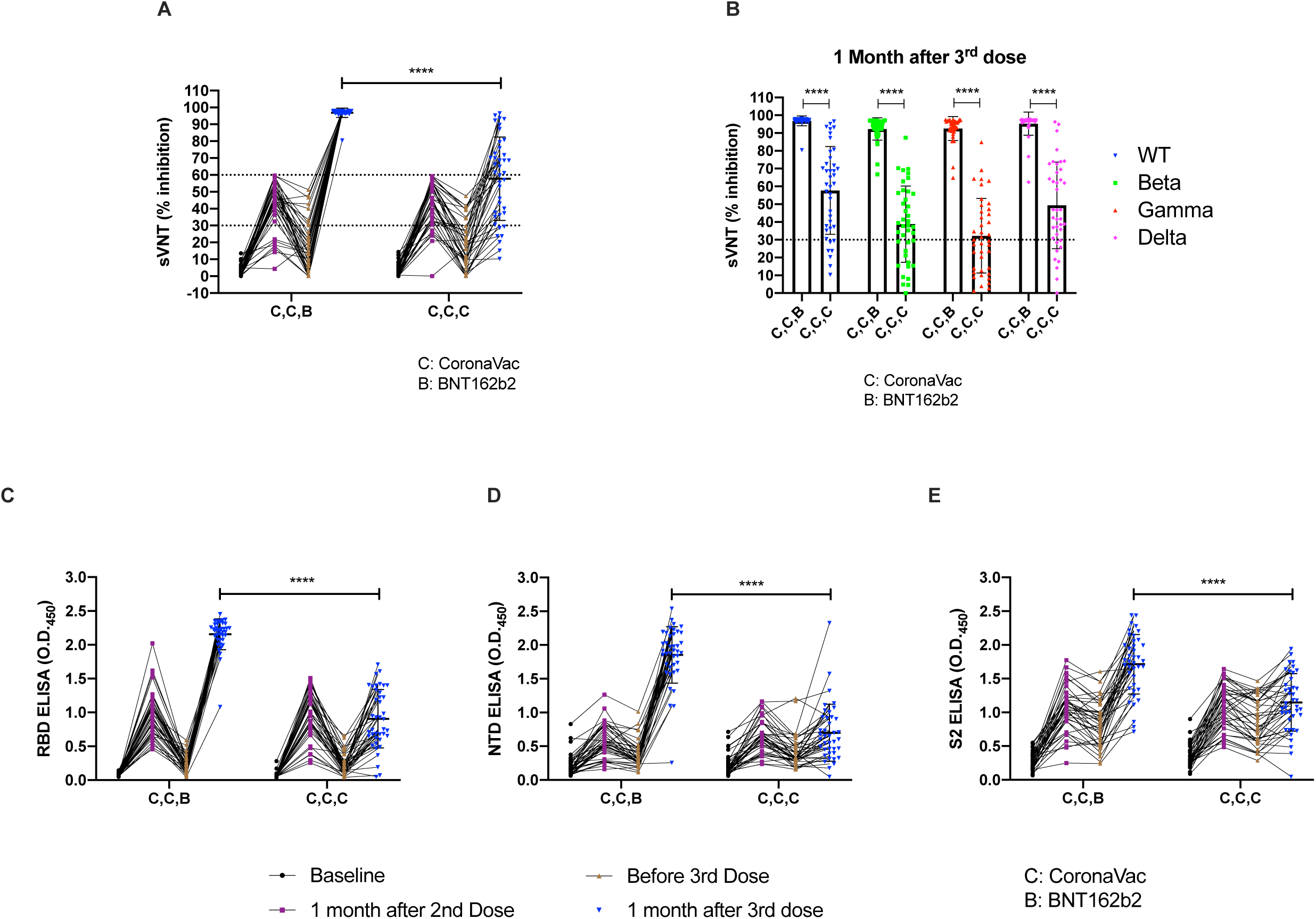
Antibody responses of individuals before and after the third dose of either BNT162b2 or CoronaVac. The levels of antibodies after the third dose of either BNT162b2 (n=40) or CoronaVac (n=40) were detected from the plasma collected from vaccinated adult individuals who had received two doses of CoronaVac. (A) The percentage of inhibition from the plasma of pre-vaccination, 1 month after two doses of CoronaVac, before and 1 month after the third dose of either BNT162b2 or CoronaVac were tested by surrogate virus neutralization test (sVNT). (B) The percentage of inhibition against the wild type, beta, gamma and delta variants was compared between the two groups at 1 month after the third dose of vaccination. The levels of (C) RBD-specific (D) NTD-specific and (E) S2-specific IgG antibodies from the plasma of pre-vaccination, 1 month after two doses of CoronaVac, before and 1 month after the third dose of either BNT162b2 or CoronaVac were tested by ELISAs. C,C,B and C,C,C indicate the vaccines (C: CoronaVac; B: BNT162b2) that were used for the first, second and third dose of vaccination. **** indicates p<0.0001

We also tested the % of inhibition from the plasma samples against different variants of concern (VOC). The results of the sVNT against the beta, gamma and delta variants in the BNT162b2 group were 92.29% (SD 6.28), 92.51% (SD 6.72) and 95.33% (SD 6.44) respectively which are significantly higher than the CoronaVac group (Beta: 38.79 % (SD 21.38), p<0.0001; Gamma: 32.22% (SD 20.95), p<0.0001;

Delta: 48.87% (SD 24.23), p<0.0001) (Figure 2B). We further compared the levels of IgG antibodies which would specifically bind to the RBD, NTD and S2 domains of the spike protein between the two groups. Similar to the results of sVNT, the plasma of participants who had received BNT162b2 showed significant higher levels of RBD (p<0.0001), NTD (p<0.0001) and S2 (p<0.0001) **s**pecific antibodies as detected by the ELISA.

## Discussion

To the best of our knowledge, this study has been the first RCT to compare the effects between BNT162b2 and CoronaVac as the third dose for vaccination. The response to the vaccine was monitored longitudinally from an age matched cohort starting from their pre-vaccination. We showed that a BNT162b2 booster elicits significantly higher neutralizing antibodies against SARS-CoV-2 including different VOC compared to using CoronaVac as a booster dose. Although more participants complained about pain and swelling at the injection site as well as fatigue and muscle pain in the BNT162b2 group compared to those receiving CoronaVac in the first week after receiving the booster dose, these were minor and no long-term adverse effect was reported by either group.

Re-infection of SARS-CoV-2 in vaccinated individuals is now a public health concern and breakthrough cases are being reported by different countries (9, 15, 16). With some vaccines, severe disease and death are also being reported in these breakthrough infections. Reasons for vaccine failure include emergence of virus variants that partially escape neutralizing antibody elicited by the wild-type virus, poor immunogenicity of the vaccine and antibody waning. Our recent observational study showed that two doses of the CoronaVac elicited significantly lower level of neutralizing antibody when compared to those who received BNT162b2 (10). We previously estimated that antibody titers of many CoronaVac vaccines would drop below the protective neutralization threshold within a few months. This contention was supported by the pre-booster vaccine sVNT levels in this study where many participants had dropped below the negative cut-off threshold of the sVNT assay prior to receiving the booster dose. Poor immunogenicity of CoronaVac was also reported by other groups (17,18).

The safety and immunogenicity of using homologous vaccine as the third dose in adults who had received with two doses of CoronaVac have been previously reported (19). This double-blind, randomized, placebo-controlled clinical trial showed that a third dose of CoronaVac administered six or more months after the second dose resulted in an increase in antibody levels. However, whether boosting with a heterologous mRNA vaccine may provide even better immunogenicity is not yet known. Two studies recently reported their results of using either BNT1626 or AZD1222 as the third dose for adults who had received two doses of CoronaVac (18,20). However, neither study was randomized and lacked longitudinal data to compare antibody levels before and after receiving the boosting dose. Importantly, adverse reactions were not evaluated in these two studies.

Our data showed that both CoronaVac and BNT162B2 vaccine boosters were safe with acceptable levels of mild adverse reactions. While both vaccines resulted in boosting neutralizing and spike binding antibody levels, BNT162b2 led to markedly higher levels of antibody comparted to those boosted with CoronaVac. Given the fact that delta variant is now dominant worldwide, our results have shown that all the participants in the BNT162b2 group achieved two times of 50% of protection threshold (estimated as 60% of inhibition in sVNT) (Supplementary Figure 1) 1 month after the booster dose in comparisons to 16 out of 40 (40%) in the CoronaVac group.

There were some limitations in our study. Firstly, the sample size in our study was relatively small, but we have shown that the large difference of the results between the two groups and provided sufficient statistical power to draw our conclusion. A larger scale RCT will be needed especially for evaluating those rare adverse effects. Secondly, our study cohort only focused on those who had poor response to the CoronaVac that were urgently in need for the third dose. In routine practice, we will need to use booster doses without serological testing to identify poor antibody responders. Finally, T cell responses were not assessed in this study. Our previous report found that CoronaVac elicited more potent T cell responses than the BNT162b2 vaccine (10). We will further study the T cell response from this cohort together with the samples collected at their six-month time point.

## Conclusion

Our RCT has shown that both CoronaVac and BNT162b2 being used as booster doses in those with poor immune responses to two doses of CoronaVac vaccine elicited increases in neutralizing antibody responses. However, BNT162b2 vaccine was markedly superior to CoronaVac when used as a booster dose. BNT162b2 not only elicited higher level of SARS-CoV-2 specific antibodies but also led to higher cross-reactive antibody levels to different VOC. The adverse reactions were mild and short-lived. In contrast, a significant proportion of those boosted with CoronaVac had insufficient (<60% inhibition in sVNT assays) neutralizing antibody responses and they had even poorer neutralizing antibody against VOC.

## Data Availability

All data produced in the present work are contained in the manuscript

## Acknowledgments

We thank Dr Fung Hong (Chinese University Medical Center), Dr Ken Tsang (Kowloon Bay community vaccination center) and Dr Beatrice Cheng (Prince of Wales Hospital) for allowing us to recruit subjects for this study. The recombinant RBD and NTD proteins were kindly provided as gifts by Prof Ian A. Wilson and Dr Meng Yuan from the Scripps Research Institute.

## Figure Legends

**Supplementary Figure 1.**
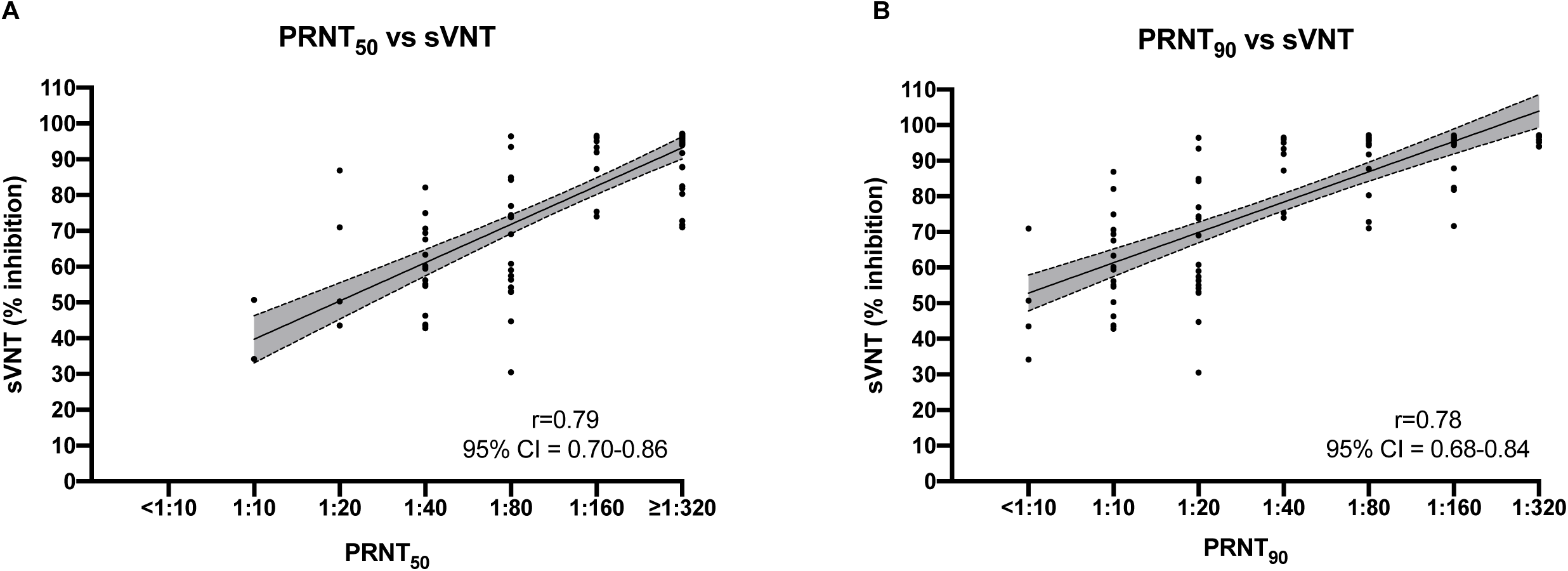
Correlation between % inhibition in surrogate neutralization tests (sVNT) and 50% plaque reduction neutralization tests (PRNT_50_) (A), PRNT_90_ from 98 samples of vaccinees (10) (B). The gray area represents the 95% confidence intervals.

